# Development and operationalization of an electronic palliative care registry in a large integrated health system

**DOI:** 10.1101/2021.05.29.21257904

**Authors:** Sylvia EK Sudat, Kathy Blanton

## Abstract

**Background:** Palliative care teams generally lack prospective tools to identify individuals who could benefit from specialty palliative care, which hinders their ability to find and treat patients early in their illness trajectories. Health systems are also limited in their ability to assess how well their palliative care services reach the population in need, which in turn makes it much more difficult to determine the quality, value, and effectiveness of those services. This study describes the creation, validation, and operationalization of an electronic registry of patients potentially eligible for palliative care at Sutter Health, a large integrated health system in northern California, US.

**Results:** The electronic palliative care eligibility algorithm performed well within the chart review validation sample, with an area under the receiver operating characteristic curve (AUROC) of 0.903 and area under the precision-recall curve (AUPRC) of 0.545. Within the 2.4 million individuals who contacted the SH electronic health record (EHR) during 2017, the final algorithm identified 1.1%-1.8% of patients (26,773-42,847) as potentially eligible for palliative care services. This included approximately 12.9%-17.7% of inpatients (16,392-22,507 out of 126,916), and 1.2%-1.9% of ambulatory care patients (20,140-32,212 out of 1.7 million).

**Conclusions:** A palliative care electronic patient registry was successfully created and operationalized. Performance based on an extensive chart review sample of Sutter Health patients indicated good capture of the palliative care-appropriate population, and this is further supported by successful identification of a majority of enrollees in Sutter Health’s palliative care programs during 2017.

## INTRODUCTION

Palliative care teams generally lack tools to prospectively identify individuals who could benefit from specialty palliative care. This hinders the ability of providers and health systems to find and treat patients as they become eligible – ideally early in their illness trajectories.^1–5^ Health systems may also unable to assess how well their palliative care services reach the population in need, as the size of that population can be difficult to quantify.^6–11^

Predictive models and other electronic solutions designed to tackle this challenge tend to rely on mortality as a proxy for palliative care eligibility. This necessitates a design that moves backward from death, and population estimates derived from such designs are not applicable to the rates of eligibility that are present in a prospective population, where people are at varying points in their end-of-life trajectories.^9,10^ Such definitions may also exclude individuals with longer prognoses who nonetheless have serious illness and could benefit from palliative care services.^11–13^ Most studies also focus on the acute setting or on specific diseases (such as cancer), and there are few examining palliative care needs in the general ambulatory patient population.^14^

This project aims to use the electronic health record (EHR) to create an electronic registry of patients potentially eligible for specialty palliative care services, agnostic to any population subset (such as payer). By highlighting the full unmet need that exists within the SH patient population, this registry could inform the design of an integrated palliative care service line which delivers on a full understanding of the volume and geography of patients in need and provides patients with more timely access to palliative care services.

The method for creating the registry builds upon previous work. As a part of the an analysis of the Advanced Illness Management (AIM) program’s impact on end-of-life resource utilization and end-of-life care quality indicators, an algorithm to identify potentially AIM-eligible individuals was developed for use with Medicare claims.^15^ This algorithm successfully identified 94% of AIM Medicare beneficiaries who died between 2010 and 2014. The algorithm was translated for use with Sutter’s EHR, and identified 67% of AIM enrollees served between January 2014 and September 2015. These studies were limited, however, by the fact that the true AIM-eligible population was not known. It was therefore impossible to compute true algorithm performance statistics, such as the sensitivity, specificity, positive predictive value, and negative predictive value, because the population of non-enrollees included patients both eligible and ineligible for AIM. The prior work also focused on deceased AIM population specifically, and was not intended to target the broader scope of all specialty palliative care in a prospective cohort.

The current study expanded the prior approach by (1) working with palliative care stakeholders to develop profiles of the patients that would ideally appear in the registry; (2) using a large chart review sample to test and assess performance of the identification algorithm; (3) applying the algorithm across Sutter Health’s patient population to obtain prospective estimated patient volumes.

## METHODS

The high-level approach to this project is displayed in Figure 1. In the initial phase, palliative care patient profiles and eligibility criteria were developed in collaboration with palliative care clinicians. Palliative care stakeholders across Sutter Health were invited to a presentation explaining the goals of the project and reviewing the patient profiles and eligibility criteria. A follow-up survey was sent electronically to those who participated, requesting feedback and approval of the presented information as a starting point for the development of an electronic patient registry. A target of 80% approval was set as a goal for this initial phase.

**Figure 1.**
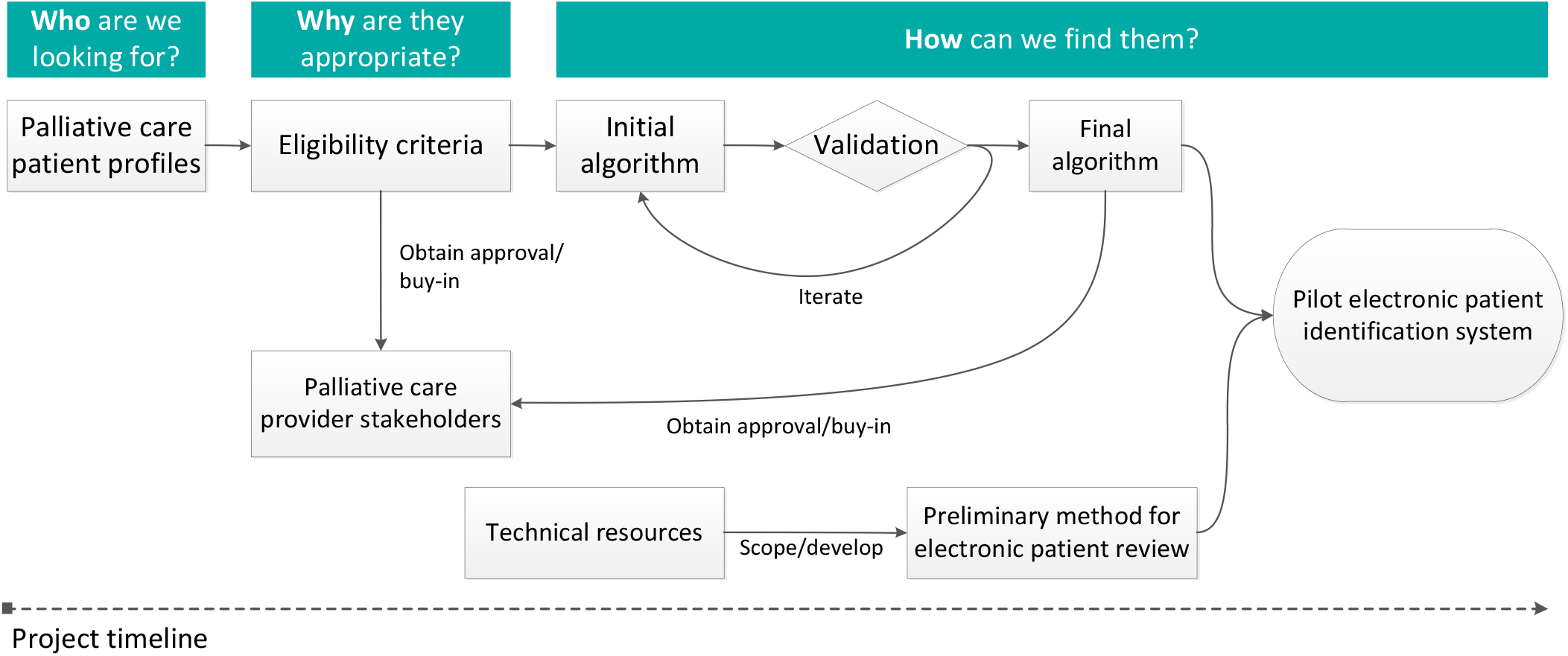
High-level project approach, assuming a progression along the timeline from left to right.

### Target population

The target population for the electronic registry (Figure 2) is the set of individuals who could potentially benefit from secondary palliative care services. We define *secondary palliative care* as advanced palliative care services delivered by specialist clinicians in a setting other than primary care, with the principal goals of comfort and quality of life. At SH, this includes the inpatient palliative care program (IPPC), Advanced Illness Management (AIM), Palliative Care Support Services (PCSS), and hospice. The palliative care-eligible patient profile, in the context of this project, is a person with a high burden of disease and one or more of the following: recent hospital utilization, complex illness management, poor prognosis, or recent decline (Figure 2).

**Figure 2.**
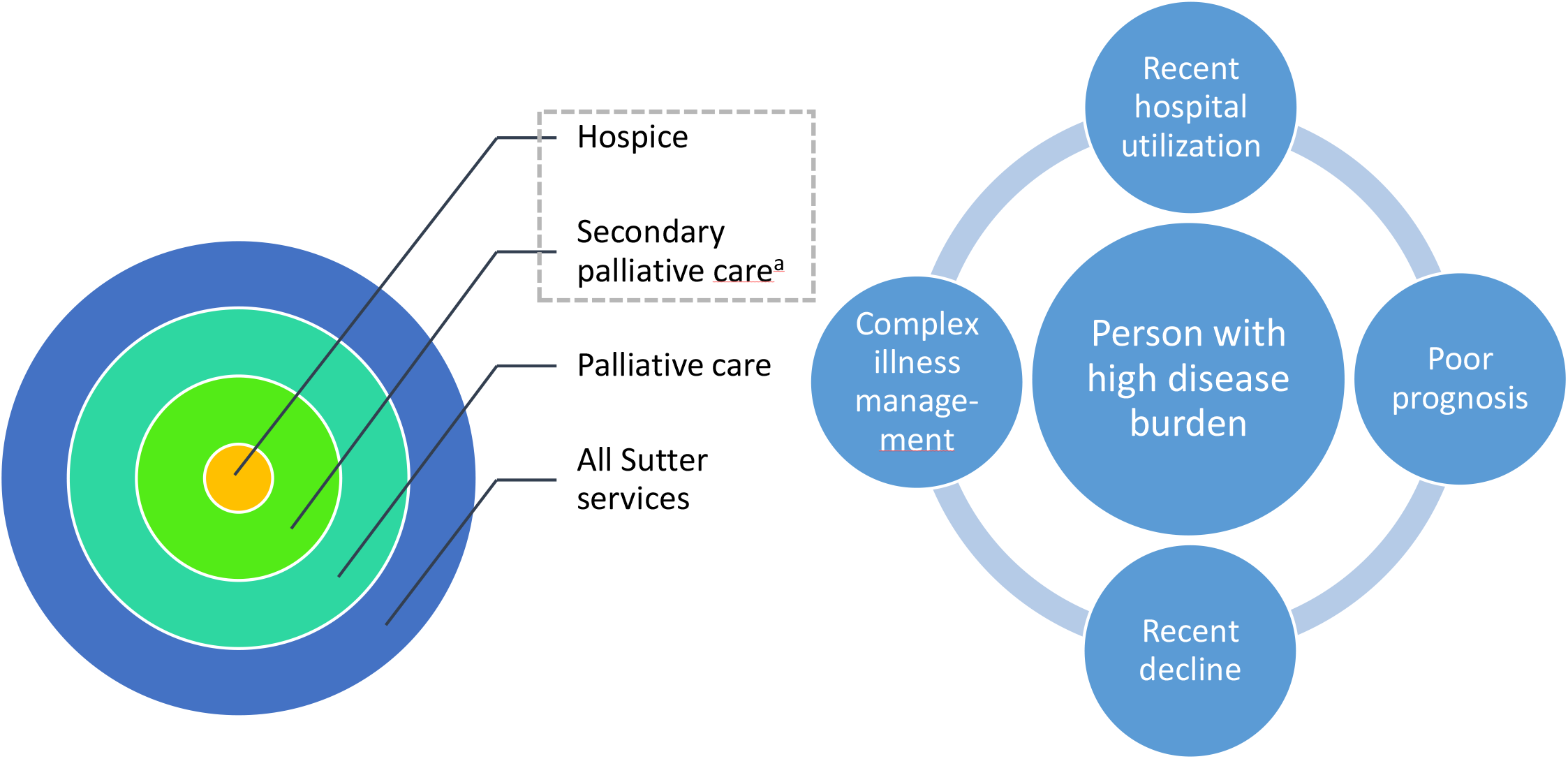
Target population for palliative care patient registry. ^a^Specialist-delivered palliative care not offered through primary care

### Chart review

In order to assess algorithm performance, design improvements, and determine the value of improvements, an extensive chart review evaluating palliative care eligibility was conducted to establish a gold standard. Chart reviews were conducted by two SH registered nurses with extensive experience receiving referrals for the AIM program. Lists of patients for chart review were created by analytical staff, and provided to the chart review nurses along with a standardized data-entry spreadsheet. For each patient to be reviewed, a reference date was pre-specified and provided on the review spreadsheet. Reviewers were asked to answer chart review questions as of the reference date, ignoring any information recorded after that date. Table 1 displays the information was requested for each chart review.

**Table 1.** Information requested for each chart review

|  |
| --- |
| Date of review |
| Does the patient have a SH foundation physician? |
| Was patient AIM eligible as of the reference date? |
| Was there enough information present in the EHR to confidently determine AIM eligibility? |
| Was patient hospice appropriate as of the reference date? |
| Was enough information present in the EHR to confidently determine hospice eligibility? |
| Was Homecare Homebase<sup>SM</sup> required to determine AIM/hospice eligibility?<sup>a</sup> |
| Was the patient enrolled in any SH palliative care program as of the reference date? |
| Additional comments related to the patient's palliative care eligibility |

<sup>a</sup>Homecare Homebase is the EHR used by Sutter's home health and hospice agency

In order to be AIM-eligible, people must have a high disease burden and meet at least one of the following four criteria: (1) rapid/significant functional/nutritional decline; (2) recurrent/unplanned hospitalizations/emergency department (ED) visits; (3) likely to die in the next 12 months (according to the referring clinician); or (4) hospice appropriate, but decline hospice services. This aligned closely with the palliative care patient definition in Figure 2.

Two chart review samples were constructed, one for each of two different reference dates: September 30, 2015 (reference date 1), and August 1, 2017 (reference date 2). Reference date 1 was chosen for practical reasons, due to the ICD-10 transition that occurred in October of 2015 (because the original algorithm code used ICD-9 diagnoses only). Reference date 2 was chosen to represent current data at the time the chart review was being conducted. The charts reviewed as of reference date 1 comprised the *training sample*, and were used to refine the algorithm. The charts reviewed as of reference date 2 comprised the *testing* or *validation sample*, and were only used to compute algorithm performance measures.

Because palliative care eligibility is a rare event, patients for chart review were selected by weighted random sampling, with oversampling of the subset of the population believed most likely to be palliative care-eligible. This included patients with evidence on the reference date of any Charlson conditions, emergency or inpatient hospital visits, or of the disease and utilization-related features shown in Table 3, rows 2-3 (ICU criterion excluded). The remainder of each chart review sample was drawn randomly from the patient population who contacted the Sutter EHR on the reference date, but were not part of the “potentially higher-risk” group. The sample sizes for the chart review are shown in Table 2.

**Table 2.** Chart review sample selection

| Reference date | Group type | Source population size | Sampled | Final chart review size |
| --- | --- | --- | --- | --- |
| September 30, 2015 | Higher risk | 6,802 | 1,114 | 2,012 |
|  | Lower risk | 70,711 | 898 |  |
| August 1, 2017 | Higher risk | 8,890 | 1,177 | 2,070 |
|  | Lower risk | 72,134 | 893 |  |

**Table 3.**
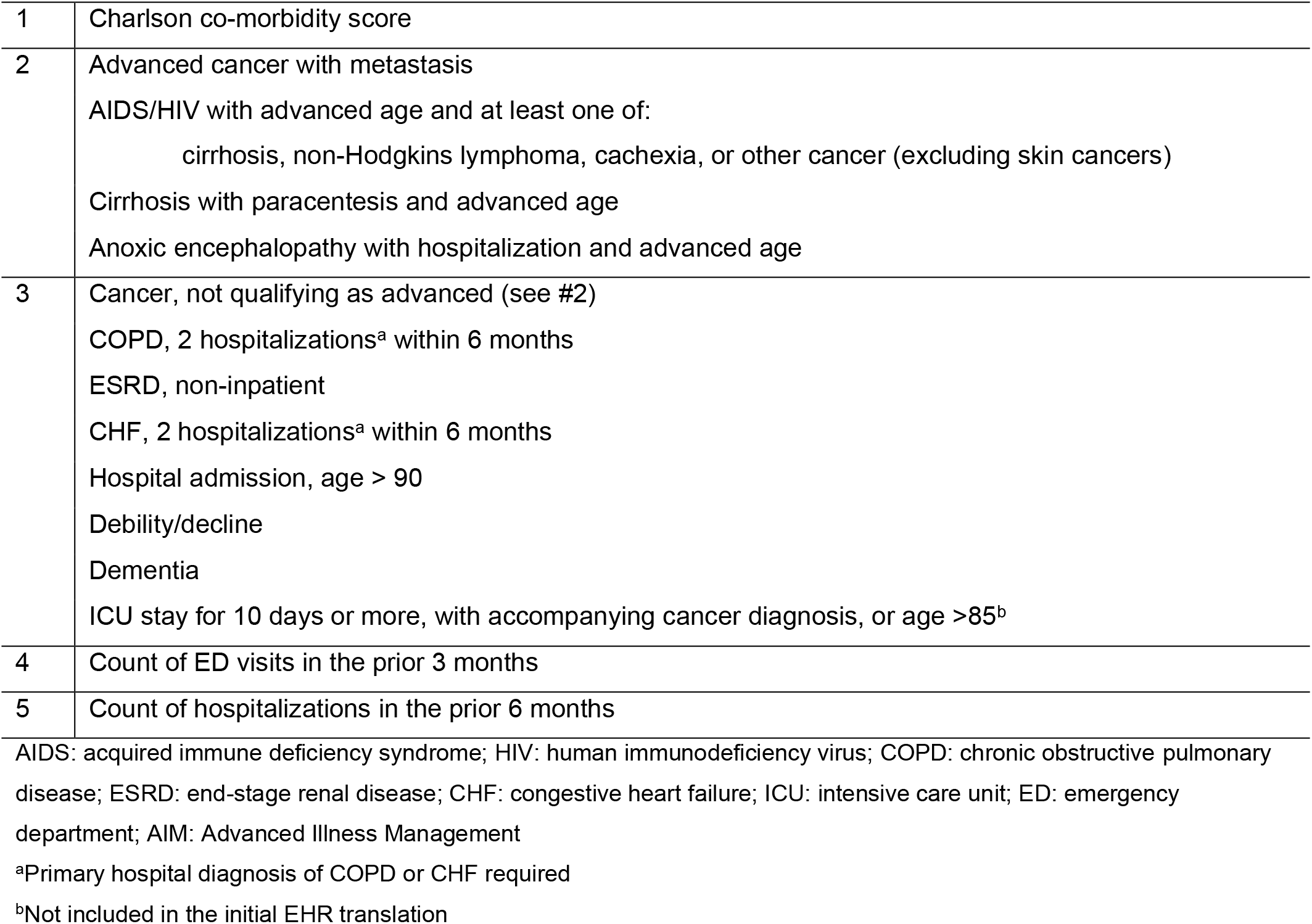
Original eligibility algorithm data domains

To maximize agreement between the two reviewers, a subset of charts were randomly chosen for assessment by both reviewers.

### Algorithm development and iteration

The starting point for creating the electronic registry was the AIM eligibility algorithm, developed in the context of the AIM evaluation for a deceased Medicare population.^15^ The algorithm utilized the data domains shown in Table 3, and was translated for use with SH EHR data in July 2017 using ICD-9 diagnoses. Initial modifications to the algorithm in the context of this project included the addition of ICD-10 codes, the implementation of the ICU criterion (see Table 3), and the additional of functional and nutritional decline criteria. Nutritional and functional decline, although part of the AIM eligibility criteria (see above), could not be readily measured from Medicare claims, and so were excluded from the original algorithm. Data from non-Sutter hospitalizations and ED visits were also included (as available) using data from Epic’s Care Everywhere®. *Care Everywhere* is the name of the health information exchange functionality built into the SH Epic EHR, which supports the secure sharing of clinical data between health care providers and organizations.^2^ Other data elements and modifications were considered in consultation with palliative care clinician stakeholders. The false positives (ineligible patients flagged as eligible by the algorithm) and the false negatives (eligible patients missed by the algorithm) in the training chart review sample were reviewed in order to gain insights into how the scoring of the data elements within the algorithm could be improved.

After the addition of each new data element or variation in algorithm scoring, the impact on performance was assessed within the chart review validation sample. Of particular importance were the sensitivity (also called recall) – the proportion of eligible patients who were correctly identified as eligible by the algorithm, and the positive predictive value (PPV, also called precision) – the proportion of patients identified by the algorithm who were truly eligible. The specificity (the proportion of ineligible patients correctly identified as ineligible by the algorithm) and the negative predictive value (NPV, the proportion of patients ineligible according to the algorithm who were actually not eligible) were also calculated.

The sensitivity, specificity, PPV, and NPV were calculated at multiple eligibility score cut-points, and relative performance between algorithm versions and score cut-points compared. Relative performance of potential algorithm improvements were also evaluated across the entire range of potential eligibility score cut-points using receiver operating characteristic (ROC) and precision-recall (PR) curves. The areas under these curves (AUROC and AUPRC) were calculated, and the Charlson co-morbidity index^16–18^ was used as a baseline for comparison. To compute the Charlson co-morbidity index, we referenced the 2005 ICD-10 and enhanced ICD-9 codes recommended by Quan et al.^4^ We used the condition weights from Quan’s 2011 update.^5,6^ A one-year (365-day) lookback period was used to calculate Charlson co-morbidity index values at each longitudinal time point using EHR data, and both hospital and ambulatory (office visit, problem list) codes were utilized. To reduce the risk of counting rule-out diagnoses, any condition that appeared only in the ambulatory or hospital outpatient setting was required to appear twice at least 30 days apart. Diagnoses that appeared in the inpatient hospital or emergency department setting were considered valid at first occurrence.

### Palliative care patient capture and volume estimates

Once the iterative algorithm development process was completed, the algorithm was run on all SH patients served in 2017, and estimates made of the size of the population potentially in need of PC services. Patients were included if they had either an EHR contact or inpatient hospital discharge in 2017. The successful identification of patients served by AIM, PCSS, or IPPC during 2017 was also computed.

## RESULTS AND DISCUSSION

### Stakeholder engagement

A total of 102 stakeholders attended the informational presentation explaining the goals of the project and reviewing the approach. Of those who attended, 33 completed the follow-up survey, and 26 (82%) approved the direction of and general approach to the project. This met the approval target of 80%.

### Chart review

A total of 4,082 charts were reviewed, 2,012 were in the training sample (9/30/2015 reference date) and 2,070 in the validation sample (8/1/2017 reference date). Of these, 5.8% were found to be eligible for AIM, and 1.7% for hospice (Table 4). There was insufficient information to determine AIM eligibility for 81 charts in the training sample (4.0%) and 68 charts in the validation sample (3.3%); hospice appropriateness could not be determined for 79 charts in the training sample (3.9%), and for 63 charts in the validation sample (3.0%). Overall, Homecare Homebase was only required 1.1% of the time (1.5% training sample, 0.6% validation sample); in approximately 96% of cases, reviewers were confident in their ability to determine AIM or hospice eligibility using data from the EHR (95% training sample, 97% validation sample).

**Table 4.** Responses to chart review questions

| Question | Percent positive response (N=4,082) |  |  |
| --- | --- | --- | --- |
|  | 2015 sample<br>(N=2,012) | 2017 sample<br>(N=2,070) | Total<br>(N=4,082) |
| Sutter foundation physician | 84.8% | 87.7% | 86.3% |
| AIM eligible | 7.1% | 4.6% | 5.8% |
| Enough data in the EHR to confidently determine AIM eligibility | 95.1% | 96.8% | 95.9% |
| Hospice appropriate | 1.8% | 1.6% | 1.7% |
| Enough data in the EHR to confidently determine hospice eligibility | 95.6% | 97.0% | 96.3% |
| HCHB required to determine AIM or hospice eligibility | 1.5% | 0.6% | 1.1% |
| Enrolled in a Sutter palliative care program | 1.3% | 1.4% | 1.4% |

A total of 236 charts were reviewed by both reviewers, and agreement across all questions was 90%. Due to staff availability, the majority of the validation sample (88%) was reviewed by a single reviewer, so a majority of the duplicate reviews (206, 87%) were conducted in the training chart review sample.

### Algorithm and eligibility score

The data elements included in the final algorithm are shown in Table 5. The AUROC for the final version in the validation sample was 0.903, and the AUPRC was 0.545, as compared to 0.891 (AUROC) and 0.414 (AUPRC) for the Charlson co-morbidity index alone. For both AUROC and AUPRC, the value would be 1.0 in the case of perfect classification. The baseline values (the values in the case of random classification) differ – for the AUROC, the baseline is always 0.5; for AUPRC, the baseline is dependent upon the prevalence of palliative care eligibility in the chart review validation sample. For this study, the AUPRC baseline is 0.05.^3^

**Table 5.** Final algorithm data domains

| Group | Element | Description |
| --- | --- | --- |
| 1<sup>a</sup> |  | Charlson co-morbidity index (CCI) |
| 2<sup>b</sup> | A | Advanced cancer with metastasis |
|  | B | AIDS/HIV disease with advanced age and at least one of:<br>cirrhosis, non-Hodgkins lymphoma, cachexia, or other<br>cancer (excluding skin cancers) |
|  | C | Cirrhosis with paracentesis and advanced age |
|  | D | Anoxic encephalopathy with hospitalization and advanced age |
| 3<sup>c</sup> | A | Cancer, not qualifying as advanced (see #2)<br>(non-melanoma skin cancers excluded) |
|  | B | COPD, 2 hospitalizations<sup>d</sup> within 6 months |
|  | C | ESRD, non-inpatient |
|  | D | CHF, 2 hospitalizations<sup>d</sup> within 6 months |
|  | E | High-risk ICU stay (duration of 3 days or more, accompanying<br>cancer diagnosis, or age >80) |
|  | F | High-risk hospital admission<br>(age > 90 or over 75 with one of: hip fracture, sepsis, delirium) |
|  | G | Debility/decline |
|  | H | Nutritional decline<br>(weight loss of 5% over 6 months, albumin < 3.0, failure to thrive, cachexia) |
|  | I | Functional decline<br>(change to dependency in any ADL over 90 days) |
|  | J | Dementia |
| 4 |  | Count of ED visits in the prior 3 months |
| 5 |  | Count of hospitalizations<sup>e</sup> in the prior 6 months |
| 6 |  | Non-compliant diabetic: Age<65; does not have cancer, AIDS, or dementia; 3 or more Charlson co-morbidities present; at least 1 ED visit in the prior 3 months OR 1 hospitalization in the prior 6 months |

AIDS: acquired immune deficiency syndrome; HIV: human immunodeficiency virus; COPD: chronic obstructive pulmonary disease; ESRD: end-stage renal disease; CHF: congestive heart failure; ICU: intensive care unit; ED: emergency department
<sup>a</sup>1-year lookback period; <sup>b</sup>6-month lookback; <sup>c</sup>60-day lookback period; <sup>d</sup>Primary hospital diagnosis of COPD or CHF required; <sup>e</sup>Short-stay non-elective admissions only

Figure 3 illustrates the tradeoff between PPV (precision) and sensitivity (recall) for 5 versions of the algorithm, with the Charlson co-morbidity index alone shown as a comparison value. Version v3.7.8 is the final version. This Figure shows how different potential improvements were compared, in order to determine the best option. It also shows how choices of different score cutoffs for determining eligibility impact the sensitivity and the PPV within the chart review validation sample.

**Figure 3.**
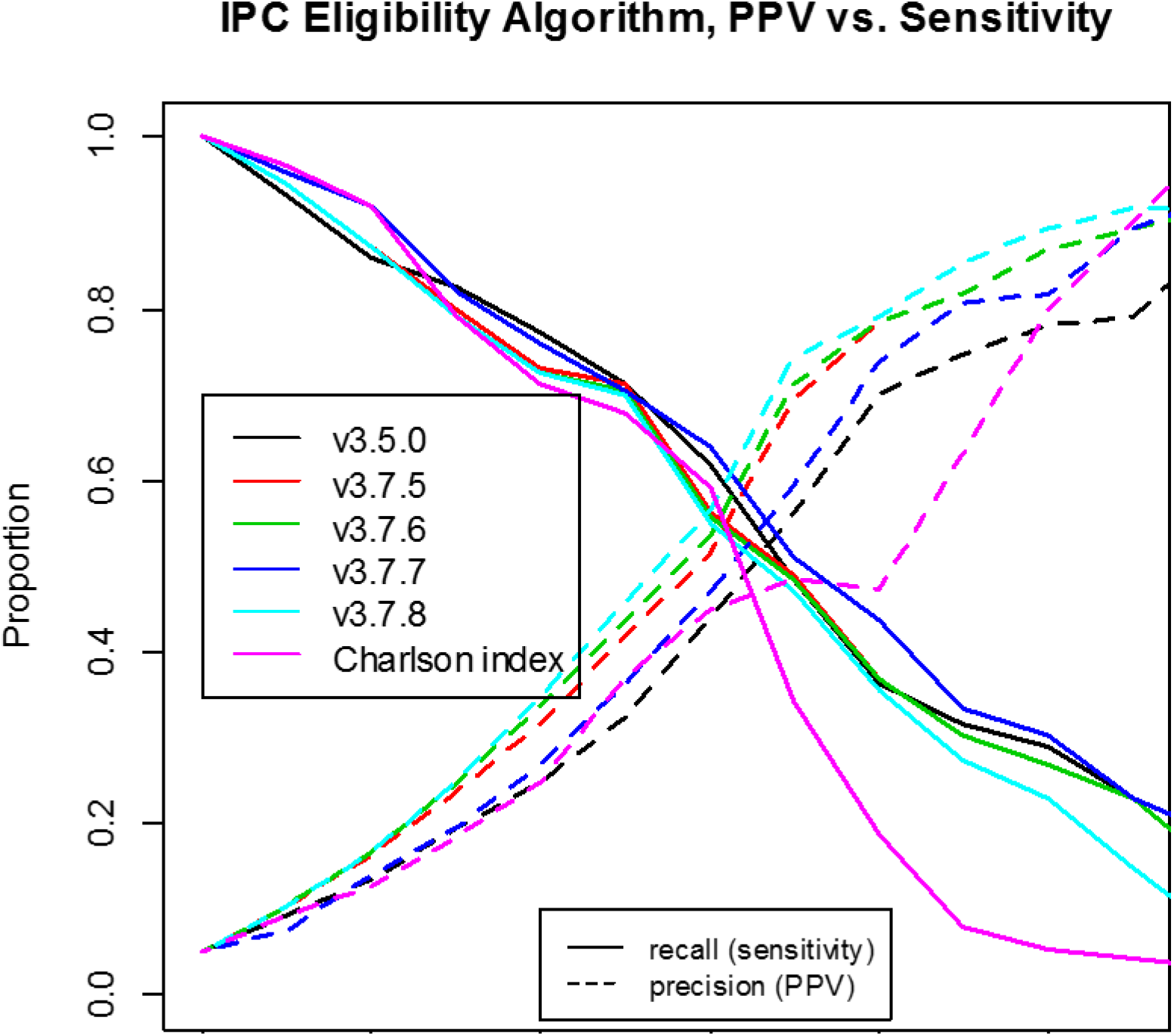
Comparison of positive predictive value (precision) and sensitivity (recall) for different algorithm versions along the range of possible values. Algorithm versions with different final digits indicate different scoring with the same data elements, and different second digits indicate different data elements. The Charlson index is included as a reference value. V3.7.8 is the final version.

Performance measures within the validation chart review sample for the algorithm at the two best eligible score cut-points are shown in Table 6. These cut-points were chosen to come as close as possible to the sensitivity/PPV break-even point, which will allow accurate estimation of the underlying eligible population size. As is expected with rare events, the algorithm is highly specific, and does not identify a large percentage of false negatives (NPV 98%, specificity 96%-98%). The sensitivity and PPV are not as high (PPV 45%-58%, sensitivity 55%-70%), which is also expected when the prevalence is low. The impact of low prevalence is also reflected in the AUROC and AUPRC values reported above.

**Table 6.** Final algorithm performance measures for two different eligible score cut-points

| Measure | Eligible score cut-point |  |
| --- | --- | --- |
|  | Option 1 | Option 2 |
| True positives | 66 | 52 |
| False positives | 81 | 39 |
| True negatives | 1826 | 1868 |
| False negatives | 29 | 43 |
| Positive predictive value (precision) | 44.9% | 57.1% |
| Negative predictive value | 98.4% | 97.7% |
| Sensitivity (recall) | 69.5% | 54.7% |
| Specificity | 95.8% | 98.0% |

### Palliative care patient capture

Table 7 shows the percent capture of patients served by AIM, PCSS, and IPPC by the algorithm during the calendar year 2017. Patients were considered correctly identified if they achieved an eligible score at any point during 2017. Based upon the choice of eligible score cut-point, 49.8%-60.4% of AIM enrollees, 17.9%-27.7% of PCSS enrollees, and 59.7%-71.6% of patients who received and inpatient palliative care consult were correctly identified as eligible by the algorithm.

**Table 7.** Percent successful identification of patients served by three Sutter Health palliative care programs in 2017

| Program | Patients served | Eligible score cut-point |  |
| --- | --- | --- | --- |
|  |  | Option 1 | Option 2 |
| Advanced Illness Management (AIM) | 6,493 | 60.4% | 49.8% |
| Palliative Care Support Services (PCSS) | 1,950 | 27.7% | 17.9% |
| Inpatient Palliative Care Consult (IPPC) | 8,205 | 71.6% | 59.7% |

The very low capture rate of PCSS enrollees can be attributed to differences in data availability based on geography – this program is only available in the areas served by Palo Alto Medical Foundation, where most hospital utilization of SH patients is at non-Sutter hospitals. Although some non-Sutter data were incorporated into the algorithm via Care Everywhere, Table 7 shows that these supplemental data were not complete enough to compensate for the data gaps.

### Estimated patient volumes

In 2017, SH served an estimated 2.44 million adult patients, including 1.71 million ambulatory care patients and 126,916 inpatients (surgical centers, birthing centers, psychiatric and rehabilitation facilities excluded). We estimate that 1.1%-1.8% of the overall population could have been palliative care-eligible (26,773-42,847), along with 1.2%-1.9% of the ambulatory care population (20,140-32,212) and 12.9%-17.7% of the hospitalized population (16,392-22,507).

### Study limitations

As with any chart review, the validity of the results is dependent upon the reliability of the reviewers. Although we did conduct dual reviews for a subset of charts and reviewed discrepancies on a regular basis with the reviewers, there could still be discrepancies in how the different reviewers assessed eligibility. The validation chart review was also largely performed by a single reviewer, so it necessarily reflects mostly the assessment of one person. The determination of appropriateness for palliative care is also not concretely defined, so is likely more subject to reviewer variability.

Because the chart reviewers were AIM intake staff, the eligibility assessment necessarily focused on AIM eligibility and not more general palliative care eligibility. Due to the reviewers’ extensive experience evaluating AIM and potential hospice appropriateness, we determined that their assessment would be most reliable if AIM-focused, rather than based upon the more comprehensive criteria developed by the palliative care stakeholder team for the SH Integrated Palliative Care initiative (with which the reviewers did not have experience). This means that patients not eligible for AIM but potentially eligible for other palliative care services would have been considered false positives, even if identified correctly by the algorithm. Future work is needed to determine where gaps in identification exist.

Finally, because this algorithm is EHR-based, it is heavily dependent upon data availability within the SH EHR, which can vary widely by geography – with large gaps in the data for those SH patients who regularly use non-Sutter facilities and providers. The portions of data from Care Everywhere used in this algorithm, while helpful additions, were not sufficient to fill these gaps. Further work with Care Everywhere data could result in more complete data capture, but is still unlikely to completely supplement the utilization data missing from the SH Epic EHR. Death data in the EHR are also very incomplete, limiting the possibility of removing deceased patients from the registry.

## Data Availability

Data are not publicly available at this time.

## ACKNOWLEDGEMENTS

The authors gratefully acknowledge the contribution of everyone on the project team to the completion of this work, most notably Anne Nilon and Julie Wilcomb, who completed the chart reviews, as well as Jeffrey Merritt (Strategy Implementation), Souvik Das (Enterprise Data Management), and Sangeeta Joshi (AIM Decision Support).

